# Little disease but lots of bites: social, urbanistic, mobility, and entomological risk factors of human exposure to *Aedes aegypti* in South Texas, U.S.

**DOI:** 10.1101/2024.02.12.24302266

**Authors:** Nicole A. Scavo, Jose G. Juarez, Luis Fernando Chaves, Nadia A. Fernandez, Ester Carbajal, Joshuah Perkin, Berlin Londono-Renteria, Gabriel L. Hamer

## Abstract

**Background:** *Aedes aegypti* presence, human-vector contact rates, and *Aedes*-borne virus transmission are highly variable through time and space. The Lower Rio Grande Valley (LRGV), Texas, is one of the few regions in the U.S. where local transmission of *Aedes*-borne viruses occurs, presenting an opportunity to evaluate social, urbanistic, entomological, and mobility-based factors that modulate human exposure to *Ae. aegypti*.

**Methodology & Principal Findings:** Mosquitoes were collected using BG-Sentinel 2 traps during November 2021 as part of an intervention trial, with knowledge, attitudes, and practices (KAP) and housing quality surveys to gather environmental and demographic data. Human blood samples were taken from individuals and a Bitemark Assay (ELISA) was conducted to quantify human antibodies to the *Ae. aegypti* Nterm-34kDa salivary peptide as a measure of human exposure to bites. In total, 64 houses were surveyed with 142 blood samples collected. More than 80% of participants had knowledge of mosquito-borne diseases and believed mosquitoes to be a health risk in their community. Our best fit generalized linear mixed effects model found four fixed effects contributed significantly to explaining the variation in exposure to *Ae. aegypti* bites: higher annual household income, younger age, larger lot area, and higher female *Ae. aegypti* abundance per trap night averaged over 5 weeks prior to human blood sampling.

**Conclusions:** Most surveyed residents recognized mosquitoes and the threat they pose to individual and public health. Urbanistic (i.e., lot size), social (i.e., income within a low-income community and age), and entomological (i.e., adult female *Ae. aegypti* abundance) factors modulate the risk of human exposure to *Ae. aegypti* bites. The use of serological biomarker assays, such as the Bitemark Assay, are valuable tools for surveillance and risk assessment of mosquito-borne disease, especially in areas like the LRGV where the transmission of target pathogens is low or intermittent.

**Author Summary:** *Aedes aegypti* is a mosquito vector with public health importance on the global scale as it transmits viruses such as dengue, chikungunya, and Zika. Although transmission rates of dengue and Zika are low in the U.S., there are a few regions, including south Texas, where local transmission has occurred. Our study aimed to evaluate the factors associated with risk of exposure to these viruses using a serological bioassay that measured antibody response to an *Ae. aegypti* salivary protein to assess human-vector contact. We collected mosquitoes, took human-blood samples, and conducted urbanistic and demographic surveys in November 2021 in eight communities in the Lower Rio Grande Valley, Texas. Our knowledge, attitude, and practices survey found that most residents recognized adult mosquitoes, though few individuals knew someone personally who been sick with a mosquito-borne disease. Outdoor adult female *Ae. aegypti* abundance was positively associated with exposure to mosquito bites. Household income, individual age, and lot area also significantly affected exposure levels. The Bitemark Assay we used in this study can be utilized as a tool for entomological risk assessment and could be used as an alternative to infection exposure in areas where mosquito-borne disease levels are low.

## Introduction

The yellow fever mosquito, *Aedes aegypti* L. (Diptera: Culicidae), is the main vector of arboviruses such as dengue, Zika, chikungunya, and Mayaro viruses. Diseases caused by these *Aedes-* borne viruses pose a threat to global health with dengue virus (DENV) affecting 390 million people annually (1), Zika having autochthonous transmission in 87 countries (2), and chikungunya causing an average annual loss 106,000 disability-adjusted life years (DALY) (3). With no effective vaccines available for protecting humans, public health authorities rely on controlling the mosquito vectors to reduce disease. *Aedes aegypti* vector control has shown variable levels of efficacy in reducing mosquito populations (4,5) and preventing disease transmission of arboviruses (6,7). With the risk of a shifting global distribution of *Ae. aegypti* mosquitoes (8) and some regions showing potential for vector presence without associated viruses (9), novel methods of surveillance and control that focus on high-risk areas are needed.

The region known as the Lower Rio Grande Valley (LRGV) along the U.S.-Mexico border in south Texas is one of the few regions in the U.S. with local vector-borne transmission of *Aedes*-borne viruses (10), and more recently human malaria (11). This can be explained by the fact that *Ae. aegypti* is well-established in the area as an efficient urban vector due to its affinity for man-made container habitat and highly anthropophilic behavior (12). *Colonias* are low-income, mostly Hispanic communities in the area that are unincorporated leading to a general lack of services (e.g., poor water sanitation) (13) and marginalized residents with little political power (14). Understanding the ecological and social factors that modulate *Ae. aegypti* abundance and human exposure to mosquito bites is key to developing efficient and effective vector control programs. Some of the previously detected risk factors for an increased indoor and outdoor abundance in low and middle-income communities of the LRGV include demographic indices (i.e., number of children and toddlers) and housing variables (i.e., air conditioning window-mounted units, number of windows) (15). Furthermore, the presence of air-conditioning reduced the risk of prior exposure to dengue virus in this region (16). However, we currently lack the understanding of how some of these risk factors relate to human-vector interactions with the added component of human mobility or how vector abundance translates to exposure to vector bites.

Current guidelines for evaluating the success of a vector control intervention recommend the use of a human outcome variable, such as active infection or past exposure to pathogens, to inform intervention efficacy (17). However, many regions with local transmission of mosquito-borne pathogens do not have consistent and sufficient burden of human disease necessary for a human disease outcome variable. For instance, West Nile virus (WNV) serosurvey in humans in Connecticut, U.S. didn’t find seropositive participants in their study despite WNV being prevalent in neighboring states and that birds and mosquitoes were infected in the region (18,19). Similarly, the LRGV represents the margin of *Aedes-* borne virus endemicity and has only sporadic local transmission of DENV (24 locally acquired cases from 2010-2017) (20). The same pattern exists for malaria transmission, with a single autochthoous human case of malaria in South Texas in 2023 (11). This makes evaluating an intervention using a human disease outcome variable difficult to utilize in *a priori* planning of a vector control intervention study.

To overcome the limitation of measuring disease transmission reduction where disease transmission is low, the use of a human antibody response to mosquito salivary proteins has emerged as a valuable tool (21–24). In this context, humans develop antibodies in response to exposure to salivary proteins associated with vector bites, and immunological assays can detect this past evidence of exposure to vector bites. Moreover, such tools can be used to estimate the risk of arbovirus exposure in areas with high transmission (25,26). It has been shown that for different mosquito species the use of IgG antibody response to salivary gland proteins can serve as effective indicators of human-vector contact as an exposure biomarker (23,27–29). Moreover, high bite exposure measured via this method has been linked to disease levels in humans for both the malaria (27,28) and dengue systems (25,30) indicating its usefulness in assessing mosquito-borne disease risk. IgG antibodies are used over IgM as their specificity is greater (25), and is specific to the genus level, with some cross-reactivity between *Ae. aegypti* and *Ae. albopictus* (31). However, immune response intensity is not linked to the likelihood of being bitten by infected mosquitoes (32). Salivary biomarkers also allow for individual mosquito bite exposure assessment and improve the ability to assess heterogeneity of disease transmission compared to community-level entomological measures (23).

Our study focuses on comparing the use of an antibody response against *Ae. aegypti* salivary gland peptides (i.e., Nterm34kDa) as an endpoint measurement in relation to mosquito abundance. Also, we aim to describe social, urbanistic, and human mobility risk factors associated with *Ae. aegypti* exposure in low-income communities (a.k.a. *colonias*) of the LRGV. The results build on our previous work in the area to elucidate seasonal patterns of mosquito abundance (33), dispersal of *Ae. aegypti* from discarded containers (34), and evaluating vector control interventions (5). Ultimately, our work can help guide public health programs to better understand the local ecology of mosquitoes along the U.S.-Mexico border and how vector control interventions might be evaluated in the LRGV and in other regions.

## Methods

### Ethic statement

This project received approval from the Institutional Review Board of Texas A&M University (IRB2021-0886D). We obtained individual written consent from each household owner for the weekly outdoor entomological surveillance and KAP surveys. We obtained individual written consent from adults that participated in the blood sampling and assent from children for the same procedure.

### Study location and site selection

The study was carried out in the county of Hidalgo, Texas, U.S., which is part of the LRGV region located along the U.S.-Mexico border. The county of Hidalgo has an estimated 870,000 inhabitants, of which 92% consider themselves Hispanic or Latino origin, 26% are foreign borne individuals and 24% live in poverty (based on income and family size/composition) (35). The climate in this region is considered humid sub-tropical, with a cold/dry season from November to February (7–21 °C), and a rainy season that starts in April (18–30 °C), peaks in September (23–33 °C) and finishes in October (19–31 °C) (36).

Sites were selected based on previous work in the area (15,33) where rapport had been built with community members. Briefly, potential sites were selected based on average income level per household, total number of households in the community, isolation of community, and distance from our base of operations in Weslaco, Texas. We selected eight low-income communities based on high community participation in past studies (Fig. 1).

**Figure 1.**
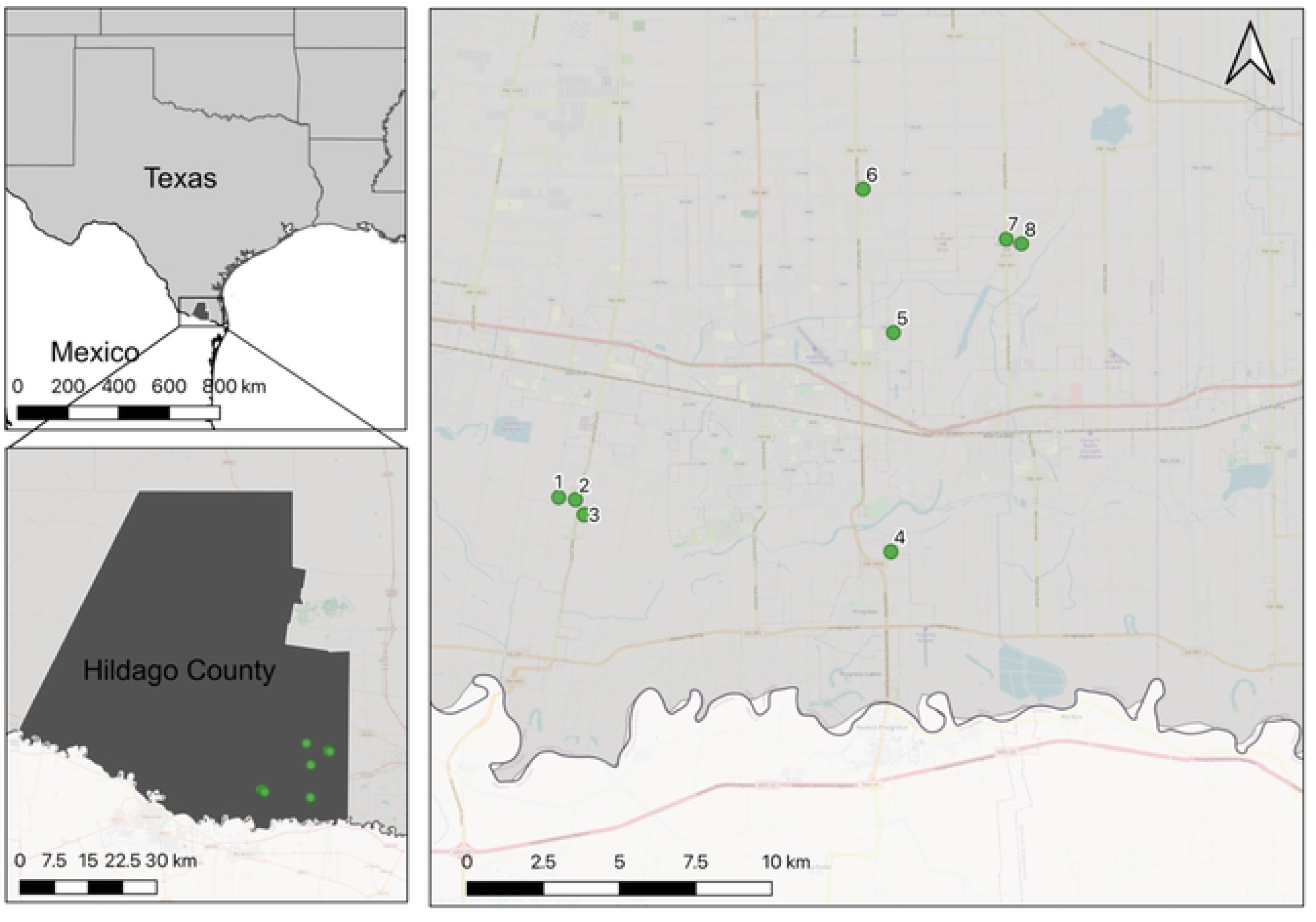
Study sites in Hidalgo county in south Texas. 1 – La Piñata, 2 – South Donna, 3 – Balli, 4 – Progresso, 5 – Chapa, 6 – Mesquite, 7 – Indian Hills West, 8 – Indian Hills East.

### Entomological Sampling

Adult mosquito sampling was carried out as part of an Auto-Dissemination Station (ADS) (BanfieldBio Inc.) intervention study. The intervention study was a cluster randomized control trial that was carried out from June 2021 until March 2022. Mosquito sampling was done using BG Sentinel 2 traps (Biogents, Germany) baited with BG lures (Biogents, Germany) placed in the peridomicile of homes at a density of 1 trap per 500m^2^. Traps were left for 24 hours once per week, collected mosquitoes were separated by sex and species (*Ae. aegypti, Ae. albopictus, Culex* sp., *Anopheles* and other) and stored at −20°C. Mosquito identification was done based on morphology using taxonomic keys (37). Households within communities were randomly selected based on desired trap density. Houses were approached for participation in the study and if homeowners agreed, a trap was placed in their lot. If a household dropped out of the study, a neighbor was recruited as a replacement in the following order: neighbor to the right, neighbor to the left, neighbor in behind, neighbor directly across the street.

### Surveys and Blood Sample Collection

Knowledge, Attitudes, and Practice (KAP) and housing quality surveys were conducted concurrently at the same household visit from November 1-13, 2021. The format of the surveys was similar to those previously done by our group in 2017 and 2018 (38). Briefly, we used a structured face-to-face questionnaire with a mixture of close-ended, semi-closed-ended, open-ended and ranking questions (see Supplementary Material 1). The KAP survey asked participants about household demographics, mosquitoes, the pathogens mosquitoes transmit, and their household member movement patterns. The house quality survey consisted of evaluating the quality of windows, doors, and their screens; housing construction material; and water holding containers and was carried out counterclockwise from the main house entrance. We recorded housing materials (timber/metal, cement, brick), screen quality (with holes, with no holes, size of holes) on windows and doors, the type of air conditioning (A/C) unit (window mounted, central) if present, and the type and number of mosquito container habitats found in each household peridomicile (see Supplementary Dataset 1). We defined the peridomicile of a household as the area between the property limit to the main house perimeter. From 95 houses under weekly entomological surveillance, 47 agreed to participate with the KAP, blood sample and housing surveys. An additional 17 houses were recruited as replacements (replacement houses were recruited from the left, right, back and front, adjacent to the BG house, if no house agreed no further house was recruited) for a total of 64 houses surveyed.

Blood sampling was done via finger prick and four circles of blood were collected on a 903-protien saver card (GE Healthcare, USA). Samples were dried and placed into a plastic bag with desiccant and stored at 4°C until further processing.

### Bitemark Assay

The *An. gambiae* gSG6-P1 peptide for the measure of exposure to *Anopheles* spp. (27) and the *Ae. aegypti* peptide Nterm-34kDa (39) were synthesized by Genscript (Piscataway, NJ, USA). ELISA conditions were standardized as described elsewhere (27,30). Briefly, dried blood samples were prepared by punching a 6mm circle out of the Whatman® 903 protein saver card (GE Healthcare, US), and eluting it into 500 µL of elution buffer (PBS 1×) and incubating overnight at 4 °C. At the time of sample preparation UltraCruz High Binding ELISA Multiwell Microplates (96-well) were coated with 100 µL/well of either gSG6-P1 or Nterm-34kDa peptide (2 μg/mL). Plates were incubated overnight at 4 °C and blocked with 200 µL of 5% skim milk solution in PBS-tween 20 (0.05%) (Blocking buffer) for 30 min at 37 °C. The sample elution was used to prepare a 1:50 dilution of the sample in blocking buffer. Then, 100 µL of that dilution were added to each well (individual samples were tested in duplicate). Plates were incubated at 37 °C for 2 h, washed three times, then incubated 1 h at 37 °C with 100 µL/well of a 1/1000 dilution of goat monoclonal anti-human IgG conjugated with horseradish peroxidase (ABCAM, Cambridge, MA). After three final washes, colorimetric development was carried out using tetra-methyl-benzidine (Abcam) as a substrate. In parallel, each assessed microplate contained in duplicate: a positive control (pool of diluted samples), a negative control (wells with no human sample), and a blank (Wells with no antigen). The blank was composed by wells containing no sample. The reaction was stopped with 0.25 N sulfuric acid, and the optical density (OD) was measured at 450 nm.

Optical density normalization and plate to plate variation was performed as previously described by our group and others (27). Briefly, antibody levels were expressed as the ΔOD value: ΔOD = ODx − ODb, where ODx represents the mean of individual OD in both antigen wells and ODb the mean of the blank wells. For each tested peptide, positive controls of each plate were averaged and divided by the average of the ODx of the positive control for each plate to obtain a normalization factor for each plate as previously described (27). Each plate normalization factor was multiplied by plate sample ΔOD to obtain normalized ΔOD that were used in statistical analyses.

### Statistical analysis

We collected 99 variables between the KAP and housing surveys. To make the analysis more manageable, we used dimension reduction methods to generate two indices (windows and doors) following the procedures described in Chaves et al. (40). To start, we carried out descriptive statistics on the KAP and housing datasets to assess which variables had low standard deviation or extremely low or high frequencies which would impact the results of our data reduction techniques. We removed variables that fell under these categories, that were colinear in nature, or that had a high degree of missing values not collected in the original surveys. If variables were colinear in nature, we kept the variables that had been shown to be important in other studies based on our knowledge of the literature. The door and window indices were generated via Principal Components Analysis (PCA) by grouping variables according to their relevance to door, window, and host categories. Other indices were tested, though only these three were kept as they explained more than 50% of the cumulative variability. Details on index creation and PCAs biplots can be found in Supplementary Material 1.

After data reduction and elimination of identification variables (e.g., street name, latitude, longitude) from the dataset, 47 explanatory variables remained. We then chose to select 12 variables from this set so that our *n*/*k* value would be above ten (41). We used our knowledge of the literature to identify 10 variables from our set that were relevant to our study (41,42). Rows with missing data were omitted from the analysis.

Our outcome variable of interest was individual exposure to *Ae. aegypti* bites measured via the Bitemark Assay (i.e., ΔOD). We analyzed how social, urbanistic, entomological, and movement factors were associated with bite exposure using a generalized linear mixed model (GLMM) approach. A mixed model was chosen to account for the potential lack of spatial independence (i.e., individuals nested within households and households nested within communities). While both linear mixed models (32,43) and GLMMs (44–46) have been used in similar serological-biomarker studies, we chose a Gaussian GLMM because they are an ideal tool for analyzing normal data whose independence is constrained by different factors (e.g., spatial or temporal) and that are modeled as random effects (47,48) in our study.

We constructed a global model (mglobal1) to evaluate the effect of 10 fixed effects on ΔOD while controlling for non-independence among houses surveyed in the same communities. The 10 fixed effected included average distance in miles traveled per week, income (2 levels: <$25,000, >25,000), host community index (host.1), door index (door.2), age (years), sex (2 levels: male, female), AC type (4 levels: window, central, mini-split, none), average abundance of *Ae. aegypti* females averaged over 5 weeks prior to sampling, area of lot (m^2^), and total containers in the lot (SM1, Table 1). Details on variable selection can be found in Supplementary Material 1. Individuals nested with homes with homes nested within each of eight communities was set as a random effect. Restricted Maximum likelihood (REML) was used due to the unbalanced nature of our dataset (47). Abundance of *Ae. aegypti* females was averaged over 5 weeks as IgG response to bites has been shown to last for a least 4 weeks (49) and is more persistent than *Anopheles* IgG responses (50).

**Table 1.**
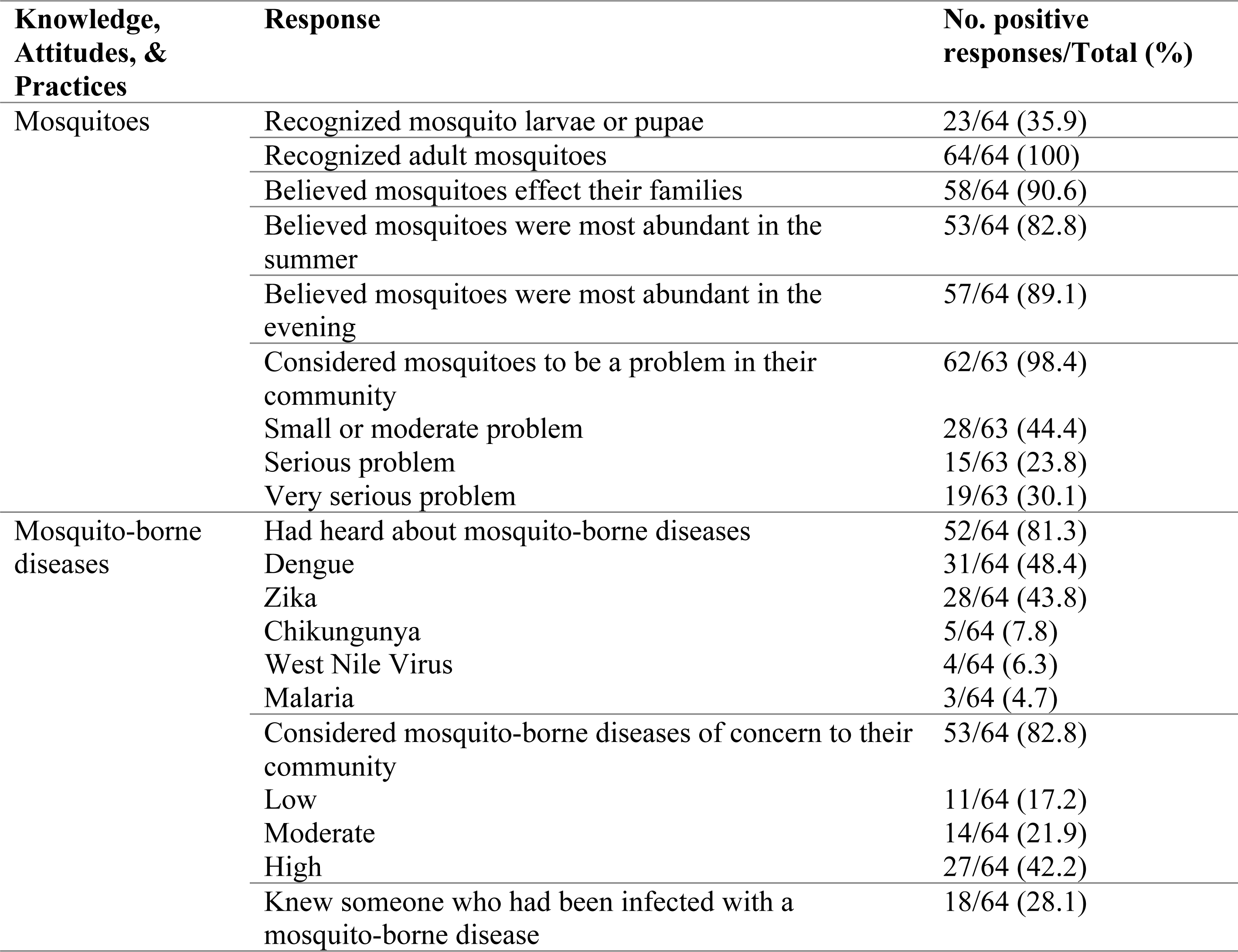
Knowledge, attitudes, and practices of household heads in the Lower Rio Grande Valley, Texas related to mosquitoes and their diseases.

We first created a global model (mgloabl1) that had a Gaussian distribution with an identity link as our outcome variable is continuous. To check model assumptions, we lotted the distribution of residuals and assessed QQ plots. Results showed deviation from the expected distribution, so we next ran another global model (mglobal2) using a log link for a Gaussian distribution. Plotted residuals from mglobal2 showed a normal distribution. Backward elimination was used to simplify mglobal2 such that simpler models with lower AIC values were kept (42,51). All models were generated, and figures were created using R Version 4.3.2 (September 1, 2023), except for Fig. 1 which was created using QGIS (version 3.16.6-Hannover). R code can be found in Supplementary Material 2 and a more detailed statistical description can be found in Supplementary Material 3.

## Results

In total, 64 adult humans from different households were interviewed using our KAP survey. The human knowledge level of adult mosquitoes was high with 100% of interviewees recognizing an adult mosquito specimen. Fewer individuals were able to identify larval or pupal mosquitoes (35.9 %). Most interviewees (90.6%) believed mosquitoes affect their families either as a nuisance, health risk, or cause of allergies, though the level of the problem they believed mosquitoes caused varied (Table 1).

Knowledge of mosquito-borne diseases was also high, 81.3% of respondents had heard of them before the interview. Most participants (82.8%) considered mosquito-borne disease a risk to their community, though few knew someone personally who had been infected (28.1%). Lots surveyed had an average size of 682 m^2^ (sd=236) with a variety of vegetation cover and vegetation height. Few lots (14.3%) actively stored water on their property for later use. However, most lots (76.6%) had other containers that could serve as larval habitat if filled with water via a rain event. Most houses had some form of air conditioning with window units being the most common followed by central systems (Table 2).

**Table 2.**
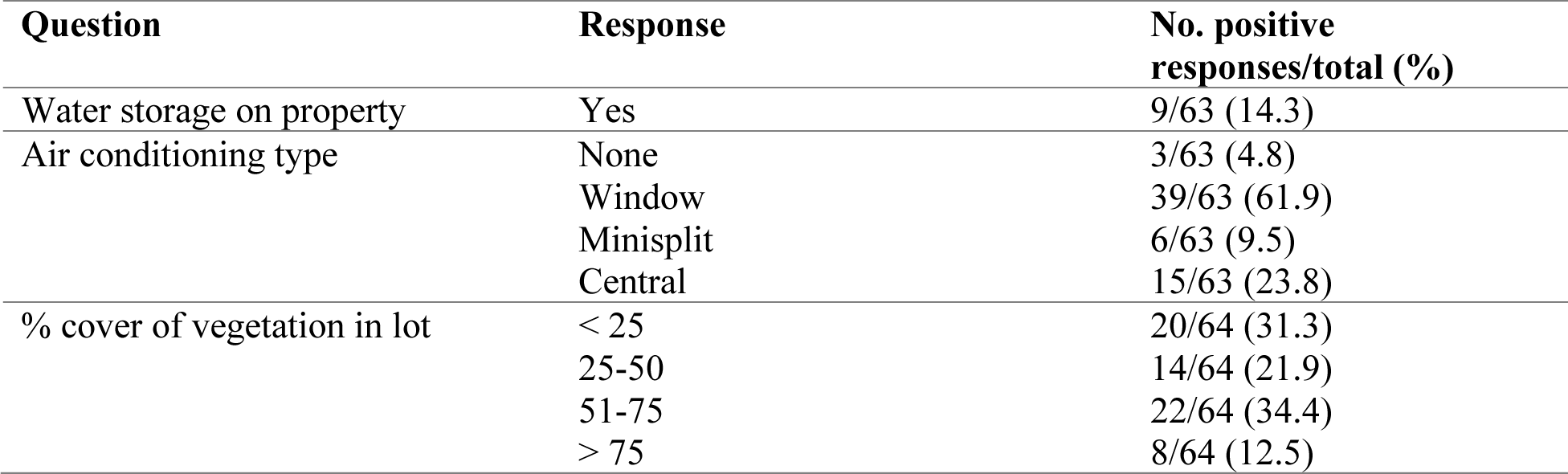
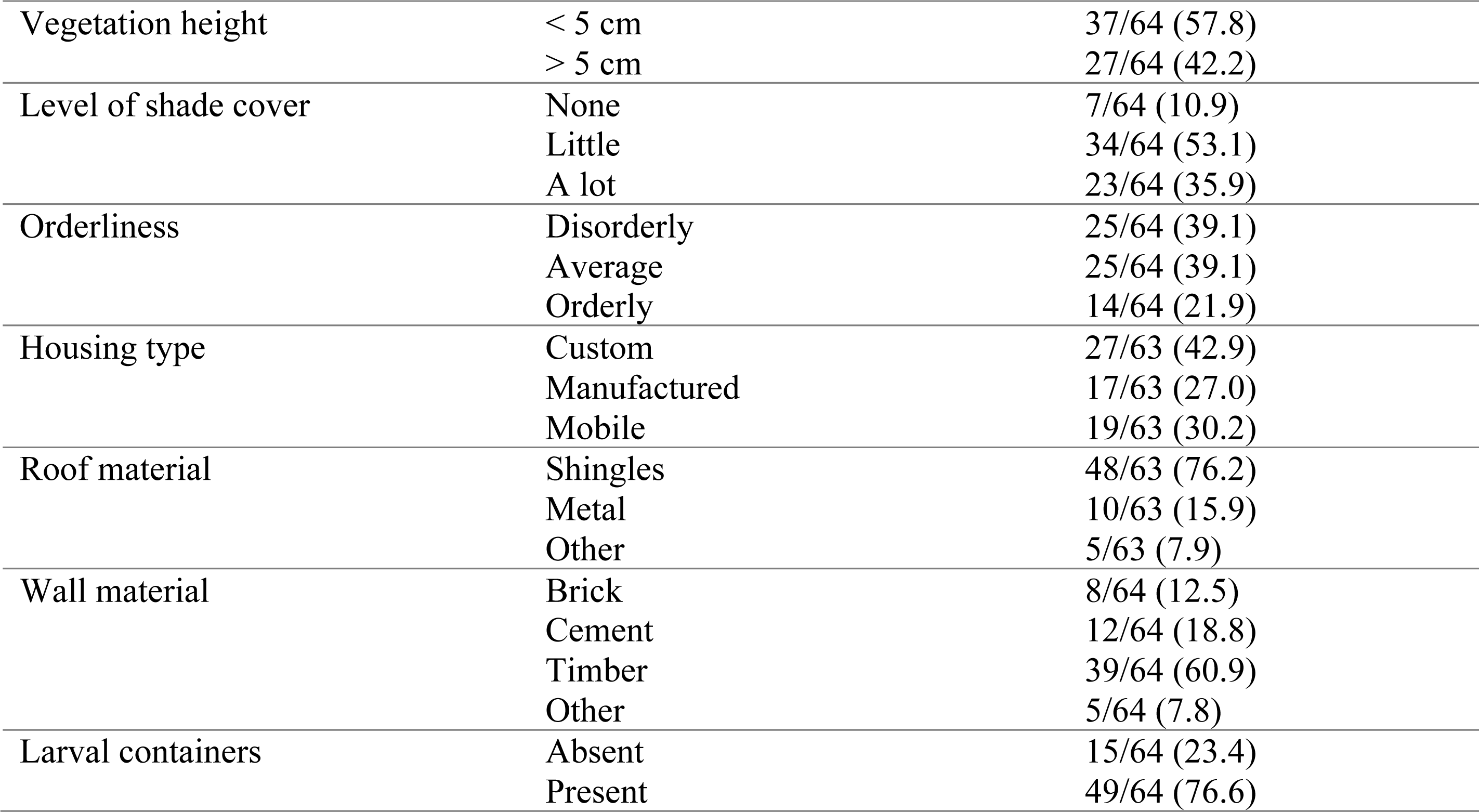
Housing and lot variables in the Lower Rio Grande Valley.

| Question | Response | No. positive responses/total (%) |
| --- | --- | --- |
| Water storage on property | Yes | 9/63 (14.3) |
| Air conditioning type | None | 3/63 (4.8) |
|  | Window | 39/63 (61.9) |
|  | Minisplit | 6/63 (9.5) |
|  | Central | 15/63 (23.8) |
| % cover of vegetation in lot | < 25 | 20/64 (31.3) |
|  | 25-50 | 14/64 (21.9) |
|  | 51-75 | 22/64 (34.4) |
|  | > 75 | 8/64 (12.5) |
| Vegetation height | < 5 cm | 37/64 (57.8) |
|  | > 5 cm | 27/64 (42.2) |
| Level of shade cover | None | 7/64 (10.9) |
|  | Little | 34/64 (53.1) |
|  | A lot | 23/64 (35.9) |
| Orderliness | Disorderly | 25/64 (39.1) |
|  | Average | 25/64 (39.1) |
|  | Orderly | 14/64 (21.9) |
| Housing type | Custom | 27/63 (42.9) |
|  | Manufactured | 17/63 (27.0) |
|  | Mobile | 19/63 (30.2) |
| Roof material | Shingles | 48/63 (76.2) |
|  | Metal | 10/63 (15.9) |
|  | Other | 5/63 (7.9) |
| Wall material | Brick | 8/64 (12.5) |
|  | Cement | 12/64 (18.8) |
|  | Timber | 39/64 (60.9) |
|  | Other | 5/64 (7.8) |
| Larval containers | Absent | 15/64 (23.4) |
|  | Present | 49/64 (76.6) |

Over the course of the five weeks before the blood samples were collected, a total of 1,379 female *Ae. aegypti* were collected, with an average of 2.9 ± 0.005 caught per trap night. The mean of ΔOD values from the Bitemark Assay for *Ae. aegypti* was 0.12, with a range of 0.05 – 0.43. No *Anopheles* were caught during the five weeks prior to human blood sampling. The range of ΔOD for *Anopheles* was 0.08 – 0.35 with a mean of 0.18 ± 0.004.

Given the large number of variables we collected, two indices were generated by grouping variables of similar nature together using PCA: door and window. Plots of the PCAs and their interpretation can be found in Supplementary Material 1. Descriptive statistics of the 12 explanatory variables can be found in Supplementary Materials 1. Minimization of AIC via backward elimination identified the best fit model which considered the following covariates: annual household income, age of participant, lot area, and the female *Ae. aegypti* abundance per trap night averaged over five weeks prior to human blood sampling. Households with an annual income of >$25,000 (i.e., 35% of households) were more 1.21(Exponentiated 95% CI: 1.06 – 1.39) times more likely to be exposed to *Ae. aegypti* bites. Age was also a significant indicator of bite exposure, with older individuals being 7% less likely to be bitten.

Larger lots were 1.11(Exponentiated 95% CI: 1.04 – 1.17) times more likely to have individuals in the household exposed to bites. For each additional adult female *Ae. aegypti* in the lot, humans were 1.12 (Exponentiated 95% CI: 1.05 – 1.18) times more likely to be exposed to bites.

## Discussion

The results from our KAP survey indicate that most residents recognized adult mosquitoes, had heard of mosquito-borne disease, and considered it to be a problem in their communities. Even so, less than one third of residents surveyed knew someone personally who had been affected by a mosquito-borne disease. These results further illustrate that the LRGV is an area of low dengue endemicity (9) where human disease outcome variables such as human dengue incidence are not good surveillance or intervention evaluation tools. In areas of low malaria endemicity where it is difficult to detect active human infection by the parasite, serological evidence of past exposure has been recommended as an alternative to measuring active infection (21,22). Moreover, one of these serological biomarkers, IgG response to species-specific salivary proteins, have been positively linked to clinical malaria (27,28) and higher viremia in dengue patients (25). Although the intensity of the immune response cannot be used as an indicator of exposure to a dengue-positive mosquito (32), the serological biomarker is a good tool to use in areas like the LRGV where infection risk is comparatively low but still present (52).

We evaluated the Bitemark Assay, which measures to IgG response to the *Ae. aegypti* Nterm-34KDa peptide, to relate female *Ae. aegypti* abundance to human exposure to their bites with the goal of evaluating an alternative tool in the surveillance of *Ae. aegypti,* their associated viruses, and interventions aimed at *Ae. aegypti* population control. Since serological biomarkers have been suggested as a cheaper and quicker option than entomological measures (52), it is important to assess the relationship between bite exposure and *Ae. aegypti* abundance. Our results corroborate a previously established link between *Ae. aegypti* abundance and bite exposure measured via a serological biomarker, a link that has been shown for both larval (44) and adult (32) abundance measures. Humans in homes with higher abundance of *Ae. aegypti* in traps had 1.12 times higher exposure for each individual mosquito than humans in homes with fewer *Ae. aegypti* in traps. We did not sample the indoor vector abundance as we have done in past studies in these communities (33), so we don’t know how this positive but weak association with outdoor abundance would have compared to indoor abundance.

Our results indicate that there are other factors – environmental and social in nature - that predict exposure to *Ae. aegypti* bites in the LRGV. Socially, age and income were significant predictors of exposure. A higher income (i.e., >$25,000), led to more exposure to *Ae. aegypti* bites, a finding that is an expected extension of Juarez et al. (2021b) work in the area who found that medium income, i.e., $25,000-50,000, was an indicator of higher outdoor *Ae. aegypti* relative abundance than in low- or high-income areas. While Martina and colleagues (33) found low-income communities to have a higher relative abundance of *Ae. aegypti* mosquitoes than mid- or high-income communities, their study design drew income data from the U.S. Census at the block level where our study and Juarez et al. (15) used income data at the household level. These differences in scale could explain the difference in the results, a phenomenon widely described in ecology as the paradox of how resource availability is described depending on a density measurement (53). Another possible explanation for these differences is that all our communities are classified as low-income and it is possible that higher income households, within this lower income context, have more exposure to *Ae. aegypti* bites. This could be due to these households have more resources for outdoor spaces such as more plant pots-saucers or water features that could serve as habitat for larva.

To the best of our knowledge, our study is the first to find a relationship between SES, as measured by income, and exposure to *Ae. aegypti* bites. Other studies have showed mixed results in terms of SES variables and bite exposure. For example, one SES variable, occupation type, has been associated with exposure previously (32) while education level has been shown to have no effect on exposure (54). In our study, age was negatively associated with exposure to *Ae. aegypti* bites indicating that younger people were more likely to be bitten than older people. In previous studies, age has been a contributor to bite exposure levels, though the directionality of the relationship varied among studies (32,44,54). Doucoure and colleagues (44) propose three hypotheses to understand the difference in exposure between adults and children: 1) antibody response is directly correlated to the bites received, 2) children have stronger reactions to bites than adults, or 3) adults experience desensitization to bites. Although this study was not designed to interrogate these three hypotheses, we observed that children had different mobility and behaviors than adults, such that they spent most of their time away from home (i.e., at school) or inside (i.e., sleeping). These differences could translate to different exposure levels to *Ae. aegypti* bites (first hypothesis). Our results showed consistency with previous work in regard to lot size (15,55), i.e., larger lots were associated with higher bite exposure.

Our study has several limitations. First, our group has been working in the LRGV in the same communities for at least 5 years. Because of this, the people we surveyed may have more knowledge about mosquitoes and their biology compared to the wider community since they have had years of exposure to our past community engagement and outreach (56). Moreover, most of our surveys were conducted during the day during the week, meaning that generally retired or individuals without normal business hour work schedules were surveyed. This could have biased the age structure of our study.

Our study supports the use of the Nterm-34kDa serological biomarker as a proxy for adult *Ae. aegypti* entomological surveillance and provides an example of how it can be effective for assessing risk in areas of low arboviral endemicity. We recommend further studies on the use of the Bitemark Assay to provide insights into the strength of this tool to measure risk factors associated with human exposure to vector bites as well as an outcome variable for vector control trials.

## Data Availability

All data produced in the present work are contained in the manuscript and supplementary materials.

## Acknowledgments

We appreciate the support of the residents and city and country public health agencies in the Lower Rio Grande Valley, TX who collaborated with us to conduct this study. We thank Danya Garza, Salvador Solis, Odaliz Sauceda, Javier Elizondo, Chris Roundy, and Charlotte Rhodes for their assistance in the field.

